# Plasma zinc status and hyperinflammatory syndrome in hospitalized COVID-19 patients: an observational study

**DOI:** 10.1101/2021.06.09.21258271

**Authors:** Gil Verschelden, Maxim Noeparast, Maryam Noparast, Maïlis Lauwers, Charlotte Michel, Frédéric Cotton, Cleo Goyvaerts, Maya Hites

**Author notes:** Shared first authors (Gil Verschelden, Maxim Noeparast).

## Abstract

Deficiency of the element zinc is associated with cytokine releasing syndrome (CRS) and the related acute respiratory distress syndrome as well as impaired antiviral response. Similar complications associate with severe SARS-CoV-2.

We conducted a prospective, single-center, observational study in a tertiary university hospital (CUB-Hopital Erasme, Brussels) to address the zinc status, the association between the plasma zinc concentration, development of CRS, and the clinical outcomes in PCR-confirmed and hospitalized COVID-19 patients. One hundred and thirty-nine eligible patients were included between May 2020 and November 2020 (median age of 65 years [IQR, 54 to 77]).

Our cohort’s mean plasma zinc concentration was 56.2 µg/dL (standard deviation [SD], 14.8) compared to 75.7 µg/dL (SD = 18.9 µg/dL) in the retrospective non-COVID-19 control group (N = 1513; *P* <.001). Markedly, the absolute majority of patients (96%) were zinc deficient (<80 µg/dL).

The mean zinc concentration was lower in patients with CRS compared to those without CRS (−5 µg/dL; 95% CI, -10.5 to 0.051; *P* = 0.048).

Among the tested outcomes, zinc concentration is significantly correlated with only the length of hospital stay (rho = -0.19; *P* = 0.022), but not with mortality or morbidity. As such, our findings do not support the role of zinc as a robust prognostic marker among hospitalized COVID-19 patients who in our cohort presented high prevalence of zinc deficiency. It might be more beneficial to explore the role of zinc as a biomarker for assessing the risk of developing a tissue-damaging CRS and predicting outcomes in patients diagnosed with COVID-19 at the early stage of the disease.

## Introduction

Severe acute respiratory syndrome coronavirus 2 (SARS-CoV-2) is associated with significant mortality and morbidity in a subgroup of patients who develop cytokine release syndrome (CRS) and the related acute respiratory distress syndrome (ARDS)^1,2^.

Zinc is the second most abundant common trace mineral in humans, yet ∼20% of the global population is estimated to suffer from different degrees of zinc deficiency^3^. Precedent evidence suggests that zinc deficiency also can be associated with lung-damaging complications such as ARDS^4,5^ as well as impaired antiviral response^3,6–9^. For instance, zinc deficiency has shown to be associated with excessive TNF-α and IL-6 activity, factors known to have a significant role in CRS^9,10^. However, the zinc status and its association with SARS-CoV-2 remains unknown.

Herein, we designed a study to determine the zinc status, explored the association between the plasma zinc concentration, the development of CRS, and the clinical outcomes in hospitalized COVID-19 patients.

## Methods

We conducted a prospective, single-center, observational study in a tertiary university hospital (CUB-Hôpital Erasme, Brussels) between May 2020 and November 2020. Consecutive hospitalized adult patients with PCR-confirmed SARS-CoV-2 infection were enrolled after obtaining informed consent. We performed clinical and laboratory assessments within 72 hours of admission. An atomic absorption spectroscopy technique was used to measure zinc concentrations in the plasma^11^.

Plasma zinc concentrations of COVID-19 patients were compared with our retrospective cohort of 1513 adult non-COVID-19 patients whose plasma zinc was previously measured in our center. The patients were selected regardless of gender or comorbidity during 2018.

The severity of COVID-19 was assessed daily during the course of hospitalization using the WHO 10-point system^12^.

Individual inflammatory parameters were assessed and summed on the day of patients’ study inclusion according to an additive six-point clinical scale, developed and validated by Webb et al.^13^, to determine the presence and severity of COVID-19-associated hyperinflammatory syndrome (cHIS)^11^. Although the authors reported an excellent discrimination^14^ of the maximum daily cHIS score for both in-hospital mortality and mechanical ventilation, external validation was still lacking. As part of this study, we independently evaluated the relevance of the cHIS scoring system and determined the best cut-offs in our cohort.

We defined the clinical outcomes as the length of hospitalization, the incidence of mechanical ventilation (including invasive and non-invasive ventilation), and mortality. We recorded the outcomes with a follow-up of 90 days from hospital admission.

### Baseline characteristics and patients’ outcomes

One hundred and thirty-nine eligible patients were included. The patient characteristics and laboratory assessments are summarized in Table and Figure 1.

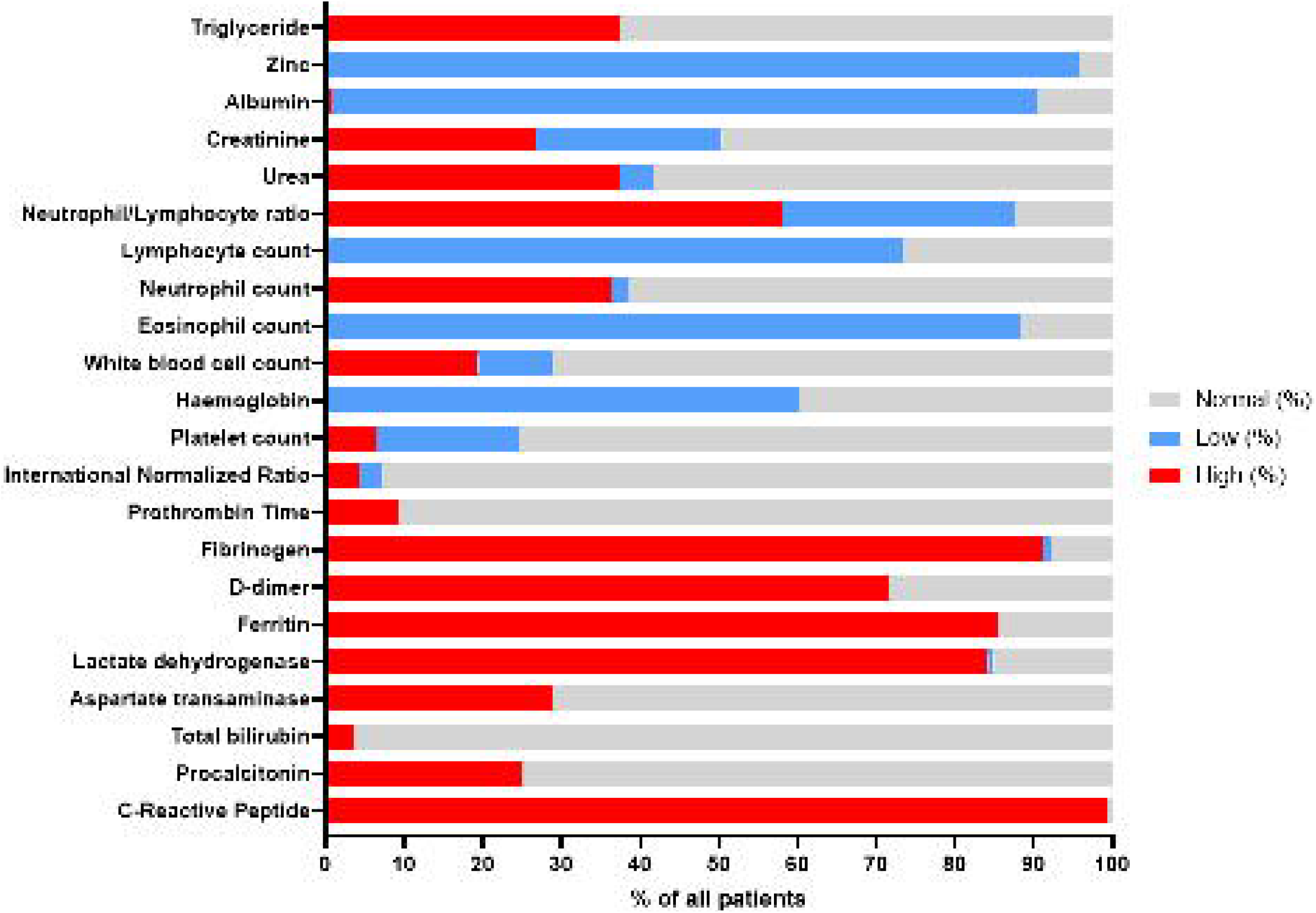

The participants were predominantly male (91/139, 65%) with a median age of 65 years (interquartile range [IQR], 54 to 77). Ninety-six patients (69%) had two or more comorbidities. The median body mass index (BMI) was 27 kg/m^2^ and 93/135 (69%) patients had a BMI >25 kg/m^2^, among which 41/135 (30%) patients were considered obese (BMI >30 kg/m^2^). The median time from symptoms onset to inclusion was eight days (IQR, 6 to 12).

The median worst daily WHO severity score was 4 (IQR, 4 to 6). Forty-two patients (30%) required mechanical ventilation or deceased during their hospitalization (WHO score >5, severe COVID-19)^10^.

Nineteen patients (14%) were admitted to the intensive care unit (ICU) during their hospital stay. Twenty patients (14%) deceased within 90 days of admission, of whom seven (35%) had been admitted to ICU.

### Zinc status and its association with patients’ outcomes

The mean plasma zinc concentration in our cohort was 56.2 µg/dL (SD = 14.8) compared to 75.7 µg/dL (SD = 18.9 µg/dL) in the control group. An unpaired t-test with unequal variance (Welch’s t-test) confirms that this difference is significant (P <.001).

Markedly, the absolute majority of COVID-19 patients (96%) were zinc deficient (<80µg/dL).

We observed that the plasma zinc concentration is weakly but significantly correlated (rho = -0.19; *P* = 0.022) with the length of hospital stay. However, the plasma zinc concentration was not significantly associated with the risk of mortality or morbidity (Table 2b). Still, the mean plasma zinc levels were systematically lower in mechanically ventilated-compared to non-mechanically ventilated COVID-19 cases (−5.23 µg/dL; 95% confidence interval [CI], -10.57 to 0.13; *P* = 0.058), and in dead compared to surviving participants (−6. 68 µg/dL; 95% CI, -13.68 to 0.33; *P* = 0.065).

**Table 1:** Characteristics of study cohort (N=139)

| Table 1: Characteristics of study cohort (N=139) | No. Available | Value |
| --- | --- | --- |
| Age (yr), median [IQR] | 139 | 65 [54-77] |
| Gender - N (%) | 139 |  |
| Female |  | 48 (34) |
| Male |  | 91 (65) |
| BMI (kg/m<sup>2</sup>), median [IQR] | 135 | 27,2 [23-48] |
| Overweight (BMI≥25) |  | 52 (38) |
| Obese (BMI≥30) |  | 41 (30) |
| Smoking status, N (%) |  |  |
| Never |  | 97 (70) |
| Former |  | 35 (25) |
| Current |  | 7 (5) |
| Number of coexisting conditions, N (%) | 139 |  |
| 0 |  | 5 (4) |
| 1 |  | 19 (14) |
| 2 |  | 19 (14) |
| >2 |  | 96 (69) |
| Coexisting conditions, N (%) | 139 |  |
| Hypertension |  | 90 (65) |
| Diabetes |  | 50 (36) |
| Hypercholesterolemia |  | 54 (39) |
| Obstructive sleep apnea syndrome |  | 11 (8) |
| Obstructive lung diseases |  | 25 (18) |
| Coronary artery disease |  | 19 (14) |
| Congestive heart failure |  | 23 (16) |
| Atrial fibrillation |  | 14 (10) |
| Chronic liver disease |  | 4 (3) |
| Chronic kidney disease |  | 31 (22) |
| Malign neoplasm |  | 9 (6) |
| Immune suppression |  | 25 (18) |
| Neurological nonvascular |  | 12 (9) |
| Stroke |  | 8 (6) |
| Days with symptoms before inclusion, median [IQR] | 131 | 8 [6-12] |
| Days of hospitalization before inclusion, median [IQR] | 139 | 1 [1-2] |
| COVID-19 medical therapy before inclusion | 139 | 108 (78) |
| Hydroxychloroquine |  | 9 (6) |
| Dexamethasone |  | 99 (71) |
| Severity of COVID-19 (worst WHO score) | 139 |  |
| Median WHO score [IQR] |  | 4 [4-6] |
| With score >5 (severe disease), N (%) |  | 42 (30) |
| cHIS score at inclusion | 134 |  |
| Median [IQR] |  | 2 [1-2] |
| With cHIS score ≥2, N (%) |  | 74 (55) |
| Plasma zinc (Normal range: [80-120µg/dL]) | 139 |  |
| Mean level (SD) |  | 56.2 (14.8) |
| With zinc deficiency, N (%) |  | 133 (96) |
| Note: IQR = interquartile range, CI = confidence interval, OR = odds ratio |  |  |

**Table 2a:** Outcomes at 90-days (N=139)

| Table 2a: Outcomes at 90-days (N=139) | Value |
| --- | --- |
| Length of hospital stay (days), median [IQR] | 8 [5-13] |
| Mechanical ventilation, N (%) | 42 (30) |
| ICU admission, N (%) | 19 (14) |
| Deceased, N (%) | 20 (14) |

**Table 2b:**
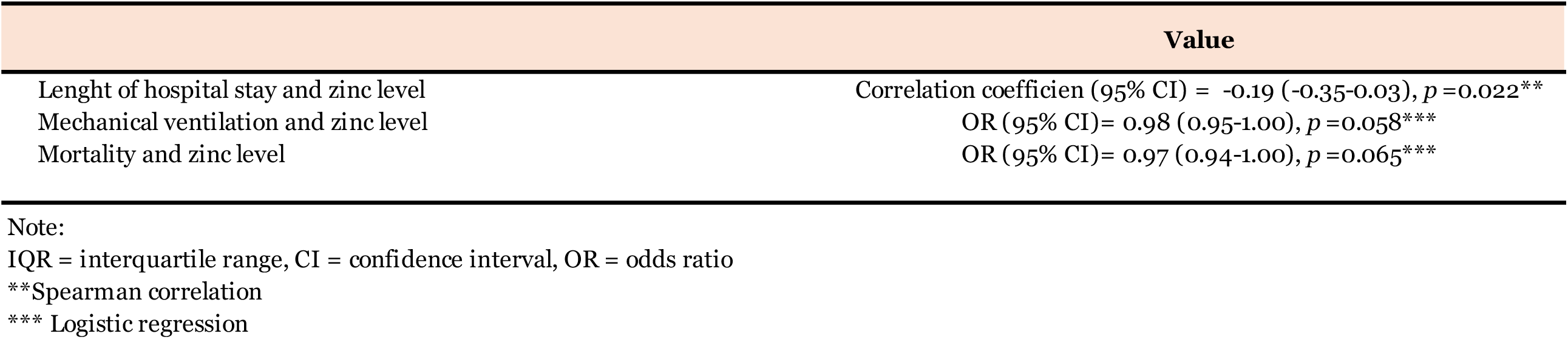
Association of outcomes and plasma zinc level (N=139)

| Table 2b: Association of outcomes and plasma zinc level (N=139) | Value |
| --- | --- |
| Lenght of hospital stay and zinc level | Correlation coefficien (95% CI) = -0.19 (-0.35-0.03), $p = 0.022^{**}$ |
| Mechanical ventilation and zinc level | OR (95% CI)= 0.98 (0.95-1.00), $p = 0.058^{***}$ |
| Mortality and zinc level | OR (95% CI)= 0.97 (0.94-1.00), $p = 0.065^{***}$ |

Note: IQR = interquartile range, CI = confidence interval, OR = odds ratio <sup>\*\*</sup>Spearman correlation <sup>\*\*\*</sup> Logistic regression

### External validation of the cHIS score as the measure of inflammatory status

The area under the receiver operating characteristic curve (AUROC) for mechanical ventilation and mortality was 0.687 and 0.674, respectively, which indicates a lack of discrimination capacity (AUC ≤0.7^14^). Nevertheless, according to the results of the Hosmer-Lemeshow goodness of fit test^14^, the score demonstrated a good calibration (p-value > 0.05), with p-values of 0.681 and 0.649 for mechanical ventilation and mortality, respectively.

Next, we aimed to identify a cHIS cut-off, which corresponds to CRS, to define a binary (low-and high-risk for poor outcome) patient group regarding inflammatory status. Our data corroborate with the cut-off suggested by Webb et al.^11^, demonstrating the best performance (Youden’s J statistic closest to one) at a threshold of ≧ 2^15^.

In our cohort, 74/134 (55%) patients had a cHIS score ≧ 2. Importantly, greater proportions of these cases needed mechanical ventilation (30/74, 40%) and died (16/74, 22%) compared to patients with cHIS scores < 2 (11/60, 18%; *P* = 0.006) and (4/60, 7%; *P* = 0.013) respectively.

Although our results support the association of CRS with both severe COVID-19 and mortality, unlike Webb et al., we were not able to demonstrate an excellent discrimination capacity of the cHIS score in our cohort^13^. In contrast to that study, most of our patients (108/139, 78%) had received immunomodulatory or suppressive therapies just before the laboratory evaluation (as part of our standard of care and based upon WHO guidelines and findings of the Recovery trial^16^). Dexamethasone reduces CRS and improves the outcome while inevitably lowers the cHIS score^16^. The cHIS score is probably more appropriate to identify high-risk patients in ambulatory practice or among the hypoxemic and dexamethasone-naïve patients.

### Association of plasma zinc concentration and cHIS score

We were not able to show a significant association between plasma zinc levels and the development of CRS (defined as cHIS score ≧ 2) in our cohort.

The mean concentration of plasma zinc was lower in patients with CRS compared to those without CRS (−5 µg/dL; 95% CI, -10.5 to 0.051; *P* = 0.048). However, with stringent and conservative Bonferroni adjustment, the results were not statistically significant.

## Discussion and limitations of the study

The small sample size and lack of a comparable control group (hospitalized for other reasons than COVID-19) are the major limitations. Additionally, as the absolute majority of the patients were zinc deficient, no room was left for any comparison between zinc deficient and non-existing zinc sufficient patients.

## Conclusions

To the best of our knowledge, we report for the first time that an absolute majority of hospitalized COVID-19 patients are zinc deficient.

We find a weak (reverse) correlation between plasma zinc concentration and the length of hospital stay, but not with mortality or morbidity.

We independently validated cHIS as a score of COVID-19 severity, but we found no significant association between zinc plasma concentration and cHIS among patients who are almost all zinc deficient.

As such, our findings do not support the role of zinc as a robust prognostic factor among hospitalized COVID-19 patients.

We recommend zinc to be measured prospectively in a larger, non-COVID-19 population to assess the incidence of the disease and CRS occurrence and its related outcomes in zinc-deficient versus zinc-sufficient individuals. We encourage further studies to explore the role of zinc as a biomarker for assessing the risk of developing a tissue-damaging CRS and predicting outcomes in patients diagnosed with COVID-19.

## Data Availability

The data that support the findings of this study are available on request from the corresponding author, (Gil Verschelden).

## Abbreviations

ARDS: Acute respiratory distress syndrome
AUROC: Area under the receiver operating characteristic curve
BMI: Body mass index
cHIS: COVID-19-associated hyperinflammatory syndrome
CI: Confidence interval
CRS: Cytokine release syndrome
ICU: Intensive care unit
IQR: Interquartile range
SARS-CoV-2: severe acute respiratory syndrome coronavirus 2
SD: Standard deviation

## Ethics approval and consent to participate

Informed consent was obtained from all participants. The study was approved by the ethics committee of Erasme Hospital, EC identifier P2020/261.

## Availability of data and materials

The datasets used and/or analysed during the current study are available from the corresponding author on reasonable request.

## Author contributions

G.V and M.Noep: conceived the idea, co-authored the research proposal, designed the study, analyzed the data, interpreted the results, generated the tables, and co-authored the manuscript. G.V: medical consultation and handling the patients’ material. M.Nop: supervised the study design and the statistical analyses and critically revised and contributed to drafting the manuscript. M.L, C.M, F.C: handled the patients’ material and performed the laboratory assessments. C.G: contributed to drafting the research proposal and study design. M.H: supervised all stages of the study, including the proposal and the study design, and critically revised and contributed to drafting the manuscript. All authors revised and approved the manuscript.

## Acknowledgments

Special thanks to Prof.Dr. Frédérique Jacobs (Université Libre de Bruxelles, Erasme Hospital).

## References

1. Leisman, D. E. et al. Cytokine elevation in severe and critical COVID-19: a rapid systematic review, meta-analysis, and comparison with other inflammatory syndromes. Lancet. Respir. Med. 8, 1233–1244 (2020).

2. Chen, L. Y. C., Hoiland, R. L., Stukas, S., Wellington, C. L. & Sekhon, M. S. Confronting the controversy: interleukin-6 and the COVID-19 cytokine storm syndrome. Eur. Respir. J. 56, 2003006 (2020).

3. Read, S. A., Obeid, S., Ahlenstiel, C. & Ahlenstiel, G. The Role of Zinc in Antiviral Immunity. Adv. Nutr. 10, 696–710 (2019).

4. Boudreault, F. et al. Zinc deficiency primes the lung for ventilator-induced injury. JCI insight 2, (2017).

5. Wessels, I. et al. Zinc supplementation ameliorates lung injury by reducing neutrophil recruitment and activity. Thorax 75, 253–261 (2020).

6. Asl, S. H., Nikfarjam, S., Majidi Zolbanin, N., Nassiri, R. & Jafari, R. Immunopharmacological perspective on zinc in SARS-CoV-2 infection. Int. Immunopharmacol. 96, 107630 (2021).

7. Noeparast, A.; Verschelden, G.; Degeyter, D.; Marco, M.; Goyvaerts, C.; Hites, M. In Light of the SARS-CoV-2 Pandemic: Revisit of the Evidence Associating Zinc and Anti-viral Response. Preprints 2020, 2020040094. In Light of the SARS-CoV-2 Pandemic: Revisit of the Evidence Associating Zinc and Anti-viral Response. Preprints 2020040094, (2020).

8. Prasad, A. S. Zinc in human health: effect of zinc on immune cells. Mol. Med. 14, 353–357 (2008).

9. Prasad, A. S. et al. Zinc supplementation decreases incidence of infections in the elderly: effect of zinc on generation of cytokines and oxidative stress. Am. J. Clin. Nutr. 85, 837–844 (2007).

10. Mehta, P. et al. COVID-19: consider cytokine storm syndromes and immunosuppression. Lancet (London, England) (2020) doi:10.1016/S0140-6736(20)30628-0.

11. Davies, I. J., Musa, M. & Dormandy, T. L. Measurements of plasma zinc. I. In health and disease. J. Clin. Pathol. 21, 359–363 (1968).

12. A minimal common outcome measure set for COVID-19 clinical research. Lancet. Infect. Dis. 20, e192–e197 (2020).

13. Webb, B. J. et al. Clinical criteria for COVID-19-associated hyperinflammatory syndrome: a cohort study. Lancet. Rheumatol. 2, e754–e763 (2020).

14. Hosmer, D. W. & Lemeshow, S. Applied Logistic Regression Second Edition. Applied Logistic Regression (2004).

15. Youden, W. J. Index for rating diagnostic tests. Cancer 3, 32–35 (1950).

16. Horby, P. et al. Dexamethasone in Hospitalized Patients with Covid-19. N. Engl. J. Med. 384, 693–704 (2021).

